# Household hunger trajectories and mental health symptoms of adolescent girls and young women in rural South Africa: an HPTN-068 longitudinal study

**DOI:** 10.64898/2026.09.01.26361988

**Authors:** Rishika Chakraborty, Molly Rosenberg, M. Margaret Weigel, Audrey Pettifor, Kathleen Kahn, F. Xavier Gómez-Olivé

## Abstract

**Purpose:** Despite the high documented prevalence of hunger and poor mental health in adolescent girls and young women (AGYW) in South Africa, this relationship remains understudied, with existing studies limited by their cross-sectional designs. This longitudinal study aimed to identify the association of hunger trajectories with anxiety and depressive symptoms, and hope in AGYW.

**Methods:** We used secondary data from the HIV Prevention Trials Network (HPTN) -068 conducted in rural Agincourt, South Africa. Complete data from 1779 AGYW collected at baseline (2011/12) and three annual follow-up visits were used. Hunger trajectories, measured using the Household Hunger Scale, were estimated via Group-Based Trajectory Modelling. Self-reported incident anxiety and depressive symptoms and hope were assessed based on AGYW’s last two follow-up visits. Covariate adjusted modified Poisson regression models estimated the association between hunger trajectories and incident anxiety symptoms, incident depressive symptoms, and hope.

**Results:** Moderate-severe hunger was prevalent in 11.0%, 10.8%, and 6.0% of the households at baseline, follow-up 1, and 2, respectively. Incident anxiety symptoms were reported by 4.5%, incident depressive symptoms by 20.0% and hopelessness by 52.8% of the AGYW. Two hunger trajectories were identified- no hunger (82%) and marginal hunger (18%). Hunger trajectories were not associated with incident anxiety symptoms [RR:1.09, 95% CI: 0.55, 2.18], incident depressive symptoms [RR: 0.97; 95% CI: 0.72, 1.33] nor hope [RR: 1.00; 95% CI: 0.81, 1.23] in AGYW.

**Conclusion:** Better understanding of the factors that promote resiliency and mental health of AGYW in this setting is warranted to inform the design of interventions.

## Introduction

Globally, nearly one in seven children between the ages of 10 and 19 years experience poor mental health [1] with the peak age of onset of mental disorders being 14.5 years [2]. Left untreated, poor mental health during adolescence can lead to short-and long-term consequences such as disrupted social relationships, academic achievements, and economic productivity, and can also progress into severe mental disorders in adult life [3, 4]. The burden of poor mental health in adolescents in sub-Saharan Africa is high, with 27%, 30%, and 41% of adolescents report having experienced depression, anxiety, and emotional and behavioral problems, respectively [5]. In South Africa, adolescents, especially girls, are reported to have a high prevalence of anxiety, depression, and post-traumatic stress [6–8].

Existing evidence suggests that household food insecurity (HFI) may be a determinant of poor mental health in adolescents [9–11]. HFI is defined as the limited or uncertain availability of nutritionally adequate and safe foods or limited or uncertain ability to acquire acceptable foods in socially acceptable ways [12]. HFI experience exists in a continuum with increasing levels of severity starting with marginal HFI, progressing to moderate and then severe, with hunger being an indicator of severe HFI [12]. In South Africa, the prevalence of severe HFI has been declining over the last 20 years [13]. Despite overall improvements in alleviating hunger in South Africa, some households continue to face challenges with accessing food. In 2018, nearly 2.1 million (11%) children lived in households that reported child hunger [14].

Relatively few studies have investigated the association of HFI with adolescent mental health in Africa [15–17]. In Ethiopia [18, 19], Kenya [20], Tanzania [21, 22], Nigeria and Ghana [19], HFI was associated with a higher risk of poor mental health in this age group. In South Africa, individuals living in food insecure hotspots had a higher incidence of depression [23]. Other studies reported that adolescents who had insufficient food in the past week had greater emotional and behavioral problems [24] and higher prevalence of psychological stress [25, 26]. However, current literature published for this geographic setting has predominantly relied on cross-sectional designs, which do not permit the establishment of temporality nor incorporate changes in HFI status over time [17]. One longitudinal study found positive associations between adverse childhood experiences (including food insecurity) and subsequent adolescent suicidal behavior [27]. Another measured HFI pre- and post-COVID onset and found it was associated with higher risk of anxiety in adolescents [28]. Notably, HFI may be transient or persistent, and different trajectories of HFI over time may impact the mental health of adolescents differently. However, it remains uncertain how changes in HFI experience over time may impact the mental health of South African adolescents.

This study aimed to assess whether trajectories of severe HFI, i.e. hunger, were associated with anxiety and depressive symptoms, and hope in school-going adolescent girls and young women (AGYW) in rural South Africa. We hypothesized that AGYW from households experiencing chronic hunger would have worse mental health symptoms compared to those from households without this experience. Our hypothesis is based on evidence indicating that AGYW from households with hunger experience emotional challenges such as worry [29], shame [30], and loneliness [31], which can harm their mental health. Additionally, AGYW experiencing hunger might need to undertake extra work for income [29], restricting their leisure and academic pursuits. Moreover, they may face a stressful home environment with suboptimal parenting [32], further affecting their mental well-being. Lastly, inadequate diet in terms of quality and quantity, which is common in households with hunger, can also have a negative impact on their mental health (Figure 1) [33]. The present study adds to the limited literature focusing on mental health and its risk and resilience factors in adolescent populations in South Africa. These findings could help to inform the design of more effective interventions to address mental health concerns of South African adolescents.

**Fig 1.**
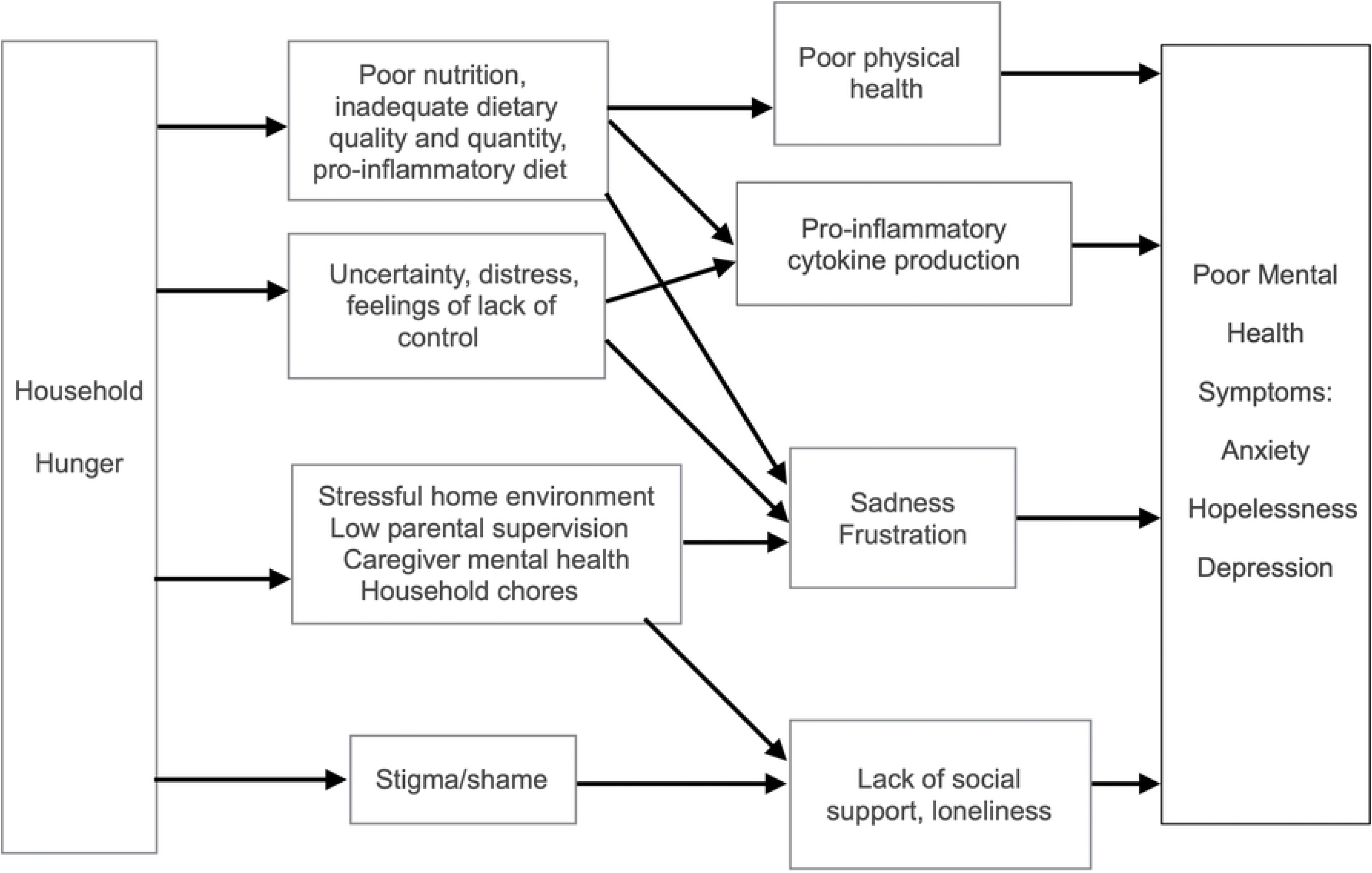
Potential pathways underlying the relationship between household hunger and mental health symptoms in adolescent girls and young women in rural South Africa

## Materials and Methods

### Study design and setting

This study used secondary data from the HIV Prevention Trials Network (HPTN)-068 study, a phase III randomized cash transfer trial, conducted in Agincourt, Mpumalanga province, South Africa [34]. The HPTN-068 study aimed to determine whether providing regular cash transfers to AGYW and their households, conditional on school attendance, reduced the risk of HIV acquisition in the AGYW [34]. The sampling frame for this study was the Agincourt Health and socio-Demographic Surveillance System (HDSS), which is run by the South African Medical Research Council/University of the Witwatersrand’s Rural Public Health and Health Transitions Research Unit [34]. The Agincourt HDSS was established in 1992 and regularly updates census data on some 20,000 households in the study area, covering a population of around 117,000 people.

For the HPTN-068, 2,533 AGYW were enrolled between March 2011 and December 2012 [34]. The HPTN-068 enrolled AGYW aged 13-20 years, enrolled in grades 8 through 11 of high school during the recruitment period, who were not married or pregnant, were able to read, lived in the Agincourt HDSS study area and intended to remain in the area until study completion, and whose caregivers possessed documentation required to open a bank account [34]. The AGYW and their households were randomly assigned to either the cash transfer or the control arm. After baseline, the AGYW were followed up annually at 12, 24 and 36 months until the study completion date or date of high school graduation, whichever came first [34]. Each young women (> 18 years) and her parents or caregivers provided written informed consent prior to study start. For AGYW under 18 years, written assent was obtained. Consent and assent forms were available in English and Shangaan. For the current longitudinal study, we used HPTN-068 data from baseline and the three waves of follow-up.

### Study participants

All AGYW enrolled in HPTN-068 who had complete data on hunger at baseline, 12-month and 24-month follow-up were eligible for inclusion in the current study. Out of 2533 participants in the parent study, 1779 (70%) had complete hunger data for all three waves (Supplementary Fig S1). This sample was used for estimating the relationship between hunger and hope. Out of 1779 participants, at baseline, 206 (11.6%) participants reported prevalent anxiety symptoms and 323 (18.2%) prevalent depressive symptoms. After exclusion of these participants and those with missing anxiety (0.6%) and depressive symptoms (3.2%) data at baseline, we created two subsamples for estimating the association of hunger with incident anxiety symptoms (n = 1563) and incident depressive symptoms (n = 1399), respectively (Supplementary Fig S1).

### Study Measures

#### Exposure- Hunger

The HPTN-068 used the 3-item Household Hunger Scale (HHS) to ask the caregivers about their experiences with hunger in the past 30 days [35]. Each question had four responses which were coded as never =0, rarely/sometimes = 1 and often = 2 [35]. The response values were aggregated to give a raw score ranging from 0 to 6, with scores 0-1 indicating little to no hunger, 2-3 indicating moderate hunger, and 4-6 indicating severe hunger [35]. Hunger trajectories were calculated using the continuous scores from the first three waves.

We used Group Based Trajectory Modelling (GBTM), a semi-parametric, finite mixture modelling approach to identify groups of households with similar trajectories of hunger [36]. Trajectory modelling was conducted in Stata/SE 17.0 (College Station, TX: StataCorp LLC. USA) using the *traj* command. We used the censored normal model [36], and specified a range of trajectory groups (two to three groups) to identify the number of trajectory groups in the data. We assumed there would be at least one hunger and one no hunger group. The shapes of each trajectory group were described using zero-order, linear and quadratic polynomial functions of time.

Final model selection to identify the appropriate number of groups and the trajectory shapes was based on the Bayesian Information Criteria (BIC) values, and other recommended diagnostic tests [36]. Each participant was assigned a trajectory group to which they had the highest posterior probability of belonging, which was our exposure. The details of the model selection process are included in Supplementary Materials Table S1.

#### Outcome – Incident Anxiety and Depressive Symptoms

The AGYW self-reported their anxiety and depressive symptoms at the time of their study exit, either at 24 months or 36 months of follow-up. We constructed incident measures for both outcomes. Anxiety symptoms were measured using the 14-item version of the Revised Children’s Manifest Anxiety Scale (RCMAS) [37]. The RCMAS has been validated in adolescents in this setting [38]. The response options were no or yes, scored as 0 or 1, respectively, and summed to produce a raw score of 0 to 14, with scores ≥10 indicating anxiety symptomatology [38]. Depressive symptoms in the past 2 weeks were measured using the 10-item Short Form of the Children’s Depression Inventory (CDI) [39], which has been validated in adolescents in this setting [26]. Each question had three response options, which were coded as 0 (most positive) to 2 (most negative), with total score ranging from 0 to 20. We used the recommended cut-off score of ≥7, the pro-rated equivalent of the full-scale cut-off, to identify depressive symptoms in the young girls [40].

#### Outcome – Hope

Self-reported hope was measured at both the 24- and 36-months follow-up visits. The HPTN-068 study team developed and validated a 12-item Hope scale in South Africa to assess the reported feelings of hope among the AGYW [41]. Each question had four response options ranging from “totally disagree” (coded as 1) to “totally agree” (coded as 4). AGYW’s responses were summed to obtain a raw score ranging from 12 to 48. We dichotomized the scores at the median to identify low (< 45) and high (≥45) hope for both follow-up waves.

#### Covariates

We used covariates including: AGYW age (in years), orphanhood (both parents alive, single orphan, double orphan), maternal education (no education, some primary or secondary education, completed secondary or higher education), study arm (cash transfer, control), asset index (quartiles), negative shocks (number of negative events), household dietary diversity score (HDDS; in numbers, higher scores indicate greater diet diversity), number of household children (in numbers), peer support (number of friends), family support (numeric score, higher scores indicate greater supportive environment), school environment (numeric score, higher scores indicate unsafe environment), and HIV status during the study period (positive or negative). Details on the measurement and construction of the negative shock variables, HDDS, HIV status measurement, family support, and school environment are described in the Supplementary Materials (Table S2) and in Table 1 footnotes.

**Table 1.**
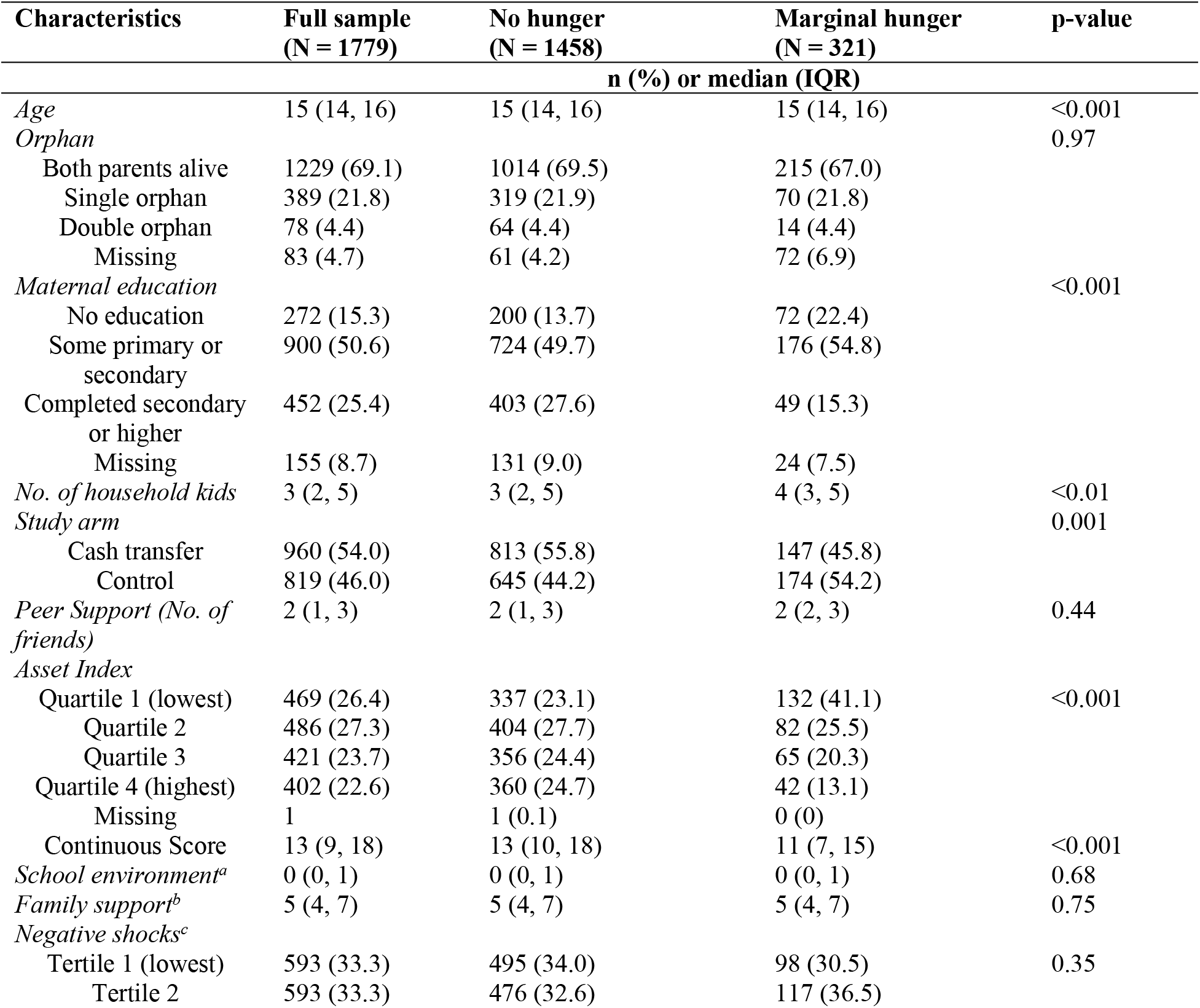
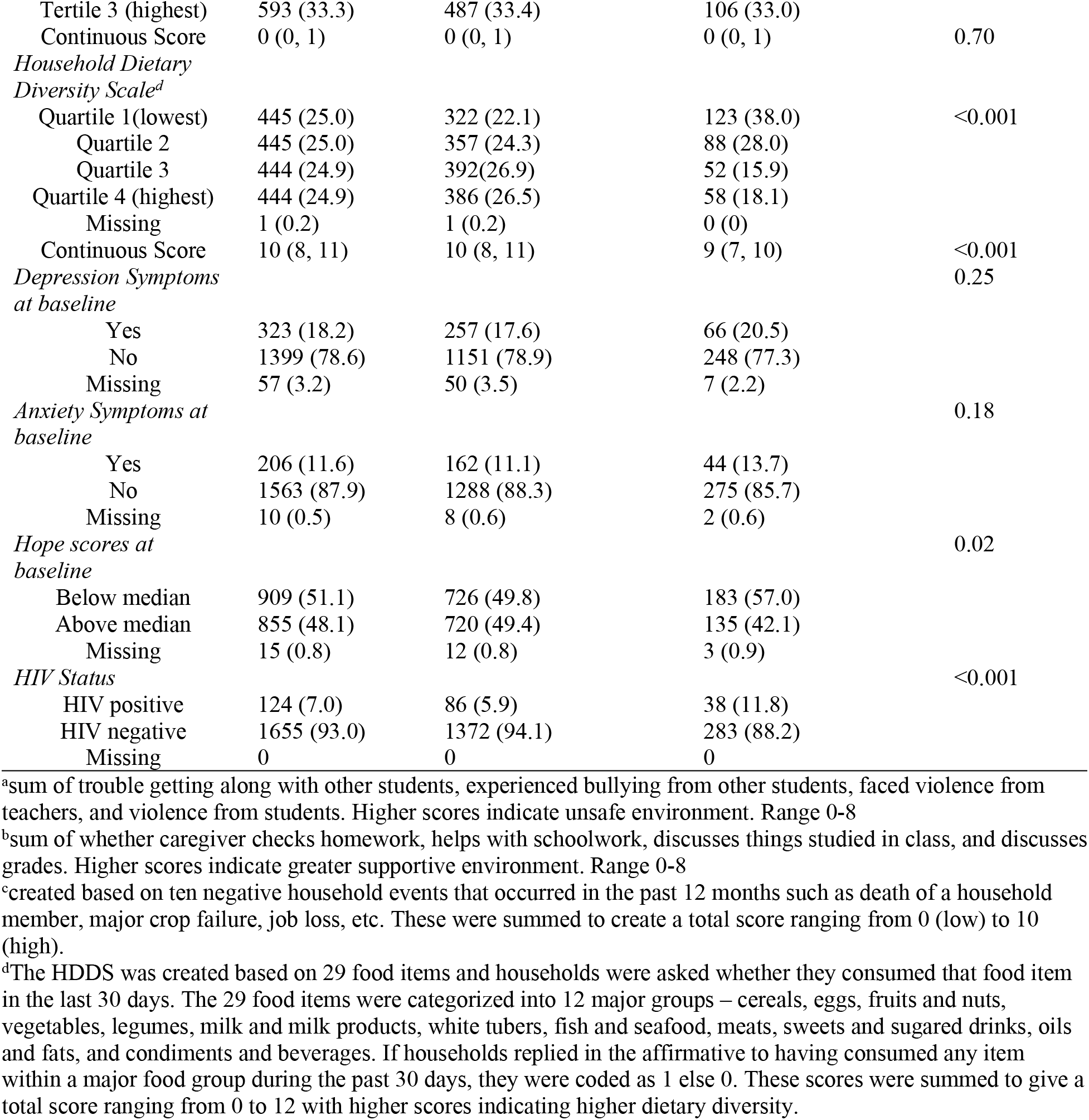
Baseline sociodemographic characteristics of adolescent girls and young women and their households according to the Hunger trajectory groups in HPTN-068 (N= 1779)

### Statistical Analysis

Statistical analyses were conducted using R statistical software (V4.3.2). The association between hunger trajectories and incident anxiety and depressive symptoms were assessed using a modified Poisson regression model with robust standard errors [42]. We controlled for waves (indicator for wave 3 or 4) in all regression models for incident anxiety and depressive symptoms to account for the different response time. In adjusted model 1, we included adolescent age, sex, maternal education, number of household children, and study arm. In adjusted model 2, we included model 1 covariates and asset index quartiles. In adjusted Model 3, in addition to model 1 and 2 covariates, we included HDDS, negative shock, and HIV status. For hunger trajectories and hope, we specified modified Poisson regressions with generalized estimating equations (exchangeable correlation structure). We also serially adjusted these models as above. We reported adjusted risk ratios (aRR) and 95% confidence intervals for all outcomes. Few AGYW (<1%) had one missing item in the RCMAS, CDI, and Hope scales. These were imputed via last value carried forward or value from the next item (Supplementary Materials for details). After imputation, complete case analysis was used.

### Sensitivity Analyses

We conducted several sensitivity analyses including a) recalculating hunger severity based on traditional HHS thresholds such that households that responded “little to no hunger” in the HHS in all three waves were classified as households with little to no hunger; households that responded “moderate” or “severe” in all three waves were classified as households with chronic moderate-severe hunger; while the rest were classified as households with transient hunger; b) using alternative thresholds for incident anxiety (cut-off at 8 and 12) and incident depressive symptoms (cut-off at 5 and 9) to test the robustness of these cut-off scores; c) using continuous scores for anxiety and depressive symptoms; d) testing interactions between hunger trajectories and HDDS, negative shock scores, and HIV status; and e) retaining those AGYW that had baseline anxiety and depressive symptoms in the model to estimate prevalence of anxiety and depressive symptoms rather than incidence. These findings are detailed in the Supplementary Analyses. Statistical significance for all hypothesis tests was set at p < 0.05.

## Results

Moderate-severe hunger was prevalent in 11.0%, 10.8%, and 6.0% of the households at baseline, follow-up year 1, and year 2, respectively. Two trajectories were identified: one group with HHS scores persistently around 0 and another with scores persistently around 1(Figure 2). Since in the HHS, scores 0-1 reflect little to no hunger [35], we categorized these two as no hunger (82.0%) and marginal hunger (18.0%) household trajectories, respectively. For both groups, the average posterior probability of belonging to each group was > 0.7, the weighted odds of correct classification were near 5, and there was sufficient proportion of samples in each group, indicating adequate model fit (Supplementary Materials Table S1). Sensitivity analyses using traditional HHS score calculations also indicated that 77% of the households experienced little to no hunger in all three waves, only 0.4% experienced moderate hunger in all three waves, while 22.6% experienced transient hunger. Notably, no household experienced severe hunger in all three waves. Given the small number (n=7, 0.4%) of households with chronic moderate hunger, we dichotomized this variable to no hunger households (77%) and transient hunger households (23%).

**Fig 2.**
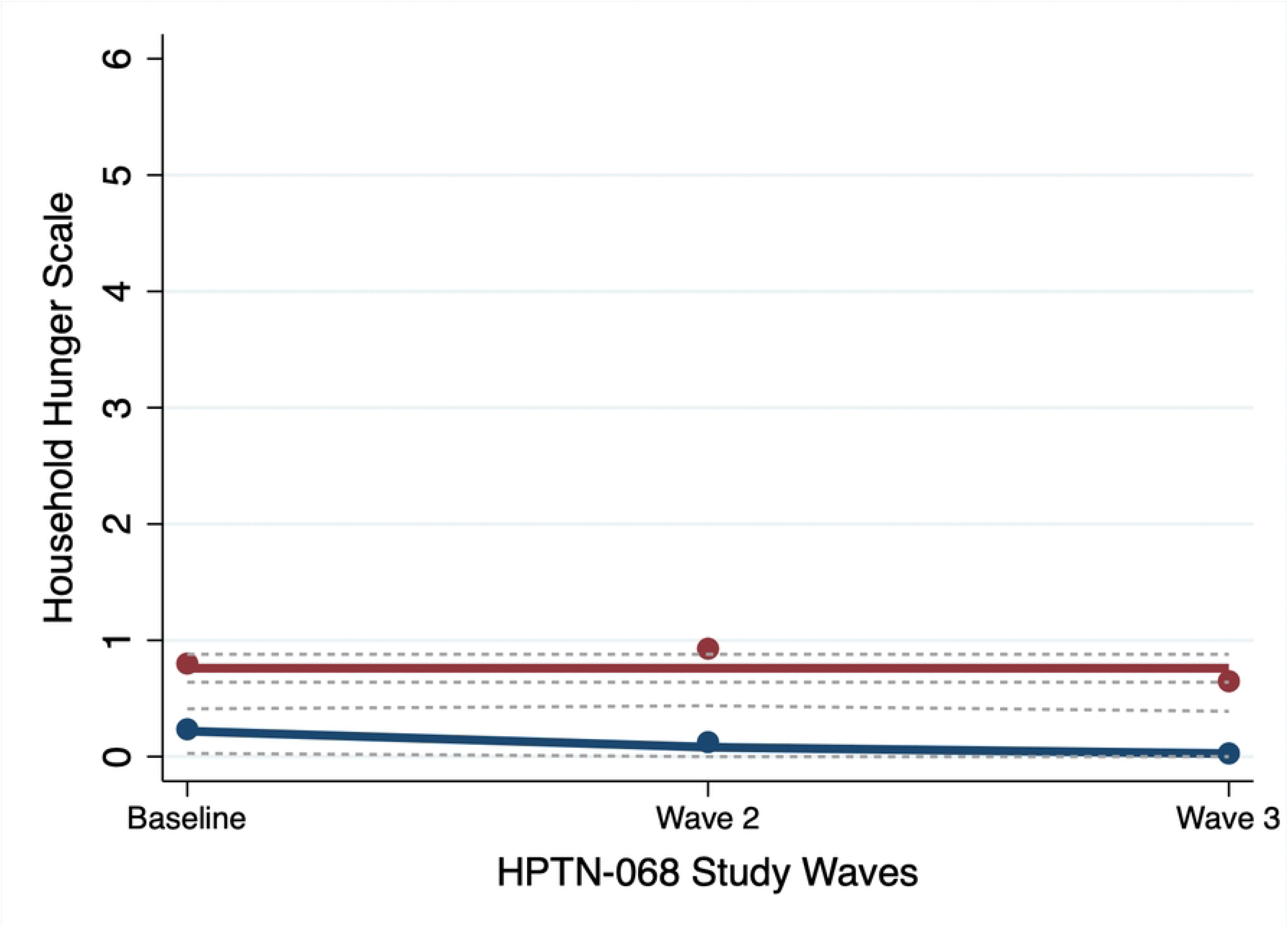
Household hunger trajectories across baseline and follow-up visits at 12 and 24 months

The AGYW in the marginal hunger households were on average slightly older (15.4 years vs. 14.9 years), were less likely to be in the cash transfer arm, and more likely to have low baseline hope scores than those from no hunger households (Table 1). Mothers from marginal hunger households were more likely to have received no education compared to their no hunger counterparts. Marginal hunger households were more likely to have a higher number of household children, be in the lowest asset index quartile, and have a lower HDDS when compared to no hunger households. Other individual- and household-level characteristics did not statistically significantly differ between no hunger and marginal hunger households (Table 1).

Incident anxiety and depressive symptoms and hope outcomes are reported in Table 2. Few AGYW reported incident anxiety symptoms (4.5%), 20.1% of the AGYW reported incident depressive symptoms, and 52.8% of the AGYW reported relatively low feelings of hope (Table 2). In the regression models (Table 2), marginal hunger household (compared to no hunger) was not associated with incident anxiety symptoms [aRR: 1.09, 95% CI: 0.55, 2.18], nor with incident depressive symptoms [aRR: 0.97, 95% CI: 0.72, 1.33], nor with hope in the AGYW [aRR: 1.00, 95% CI: 0.81, 1.23]. Similar to the trajectory modelling findings, using traditional HHS thresholds also showed that transient hunger households (compared to no hunger) was not associated with incident anxiety symptoms [adjusted RR: 0.89, 95% CI: 0.44, 1.81], nor with incident depressive symptoms [adjusted RR: 0.98, 95% CI: 0.73, 1.31], nor with hope in the AGYW [adjusted RR: 0.99, 95% CI: 0.84, 1.19] (Supplementary Tables S3-S5).

**Table 2.**
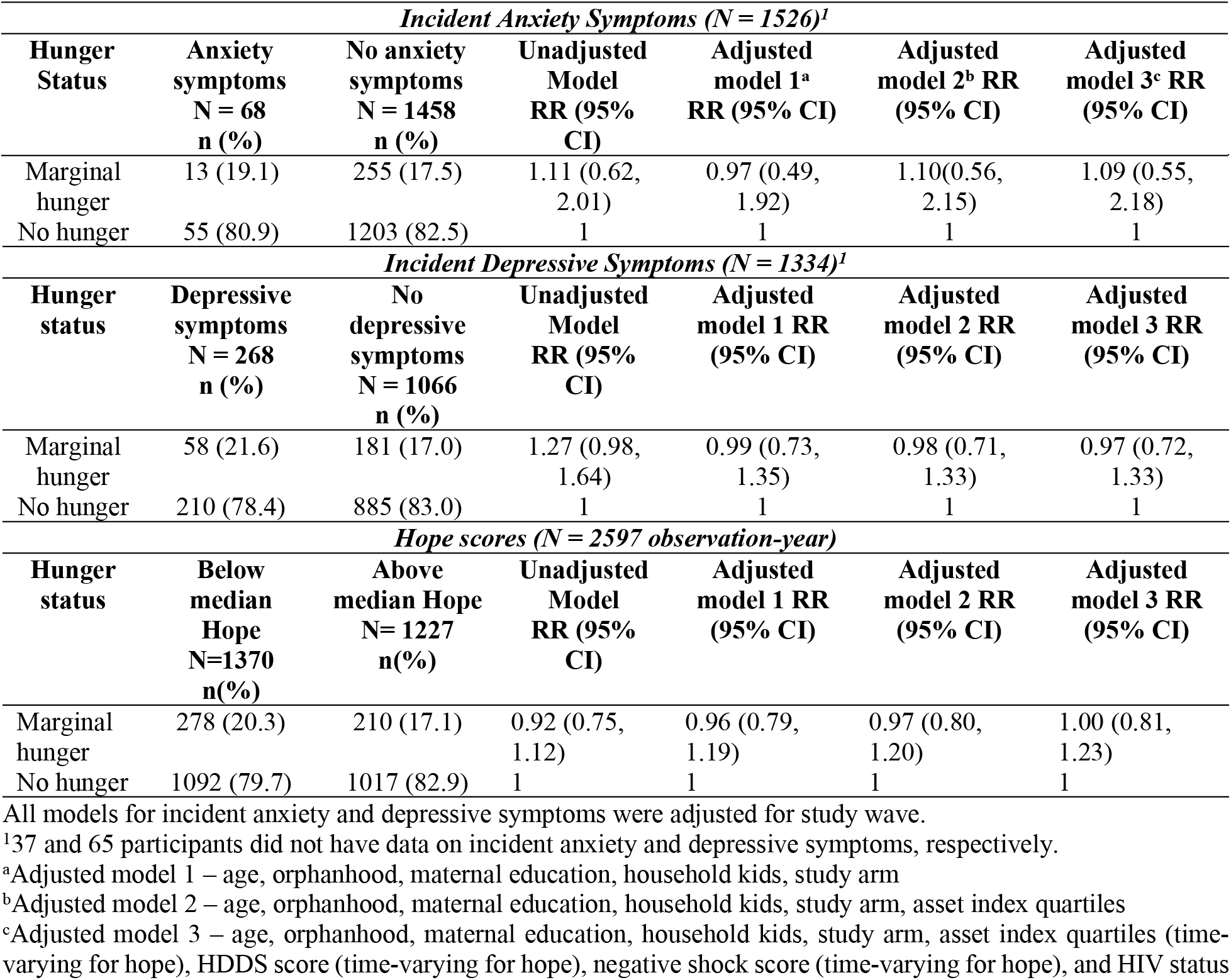
Hunger trajectories and mental health symptoms in adolescent girls and young women in Agincourt enrolled in HPTN-068 (2011/12-2015/16)

| *Incident Anxiety Symptoms (N = 1526)<sup>1</sup>* |  |  |  |  |  |  |
| --- | --- | --- | --- | --- | --- | --- |
| Hunger Status | Anxiety symptoms<br>N = 68<br>n (%) | No anxiety symptoms<br>N = 1458<br>n (%) | Unadjusted Model<br>RR (95% CI) | Adjusted model 1<sup>a</sup><br>RR (95% CI) | Adjusted model 2<sup>b</sup> RR<br>(95% CI) | Adjusted model 3<sup>c</sup> RR<br>(95% CI) |
| Marginal hunger | 13 (19.1) | 255 (17.5) | 1.11 (0.62, 2.01) | 0.97 (0.49, 1.92) | 1.10(0.56, 2.15) | 1.09 (0.55, 2.18) |
| No hunger | 55 (80.9) | 1203 (82.5) | 1 | 1 | 1 | 1 |
| *Incident Depressive Symptoms (N = 1334)<sup>1</sup>* |  |  |  |  |  |  |
| **Hunger status** | **Depressive symptoms**<br>**N = 268**<br>**n (%)** | **No depressive symptoms**<br>**N = 1066**<br>**n (%)** | **Unadjusted Model**<br>**RR (95% CI)** | **Adjusted model 1 RR (95% CI)** | **Adjusted model 2 RR (95% CI)** | **Adjusted model 3 RR (95% CI)** |
| Marginal hunger | 58 (21.6) | 181 (17.0) | 1.27 (0.98, 1.64) | 0.99 (0.73, 1.35) | 0.98 (0.71, 1.33) | 0.97 (0.72, 1.33) |
| No hunger | 210 (78.4) | 885 (83.0) | 1 | 1 | 1 | 1 |
| *Hope scores (N = 2597 observation-year)* |  |  |  |  |  |  |
| **Hunger status** | **Below median Hope**<br>**N=1370**<br>**n(%)** | **Above median Hope**<br>**N= 1227**<br>**n(%)** | **Unadjusted Model**<br>**RR (95% CI)** | **Adjusted model 1 RR (95% CI)** | **Adjusted model 2 RR (95% CI)** | **Adjusted model 3 RR (95% CI)** |
| Marginal hunger | 278 (20.3) | 210 (17.1) | 0.92 (0.75, 1.12) | 0.96 (0.79, 1.19) | 0.97 (0.80, 1.20) | 1.00 (0.81, 1.23) |
| No hunger | 1092 (79.7) | 1017 (82.9) | 1 | 1 | 1 | 1 |

All models for incident anxiety and depressive symptoms were adjusted for study wave.
<sup>1</sup>37 and 65 participants did not have data on incident anxiety and depressive symptoms, respectively.
<sup>a</sup>Adjusted model 1 – age, orphanhood, maternal education, household kids, study arm
<sup>b</sup>Adjusted model 2 – age, orphanhood, maternal education, household kids, study arm, asset index quartiles
<sup>c</sup>Adjusted model 3 – age, orphanhood, maternal education, household kids, study arm, asset index quartiles (time-varying for hope), HDDS score (time-varying for hope), negative shock score (time-varying for hope), and HIV status

Results from other sensitivity analyses indicate our findings are robust to different specifications of the outcomes. We re-ran the analyses using continuous scores for incident anxiety and depressive symptoms, and hope scores and found no associations between hunger trajectories and these outcomes (Tables S6-S8). Our findings were stable even when using alternate cut-off scores for incident anxiety and depressive symptoms (Tables S9-S12), or for prevalent anxiety and depressive symptoms (Tables S13-14). Moreover, there was no significant effect modification by HDDS, negative shocks, nor HIV status for this association (Tables S15-S16).

## Discussion

This study found no association between household hunger trajectories and incident anxiety and depressive symptoms, and hope in this sample of AGYW in Agincourt, between 2011-2016. Notably, nearly 80% of the households experienced no hunger across all waves. However, about one in five households experienced marginal hunger. Our findings suggest that while marginal hunger continues to be a burden in some study households with AGYW in Agincourt, it was not associated with mental health symptoms in the study participants.

Our findings contrast with prior studies from South Africa that reported a positive association between hunger and poor mental health [26, 28, 43]. However, two longitudinal studies reported null associations between chronic HFI and anxiety and depressive symptoms in U.S. adolescents [44, 45]. One potential reason for this inconsistency may be due to the differences in food insecurity measurement. Some studies used a single question asking whether the children had enough food to eat in the past week [25–27], others asked how often there was insufficient food in the household [43], or whether there was adequate household food in the past month [23], or used the Household Food Insecurity Access Scale (HFIAS) [28]. The present study used the HHS, which identifies the most severe experiences of HFI, specifically focusing on hunger. Thus, it is possible that households identified with “no hunger” may have experienced worry/anxiety about HFI or reduced their quality of food, which the HHS scale is unable to measure. Therefore, findings from this study are not directly comparable with other studies that used different metrics to assess both child-level and household-level food insecurity.

Importantly, we found a relatively low prevalence of hunger in this sample, which aligns with prior recent evidence from this rural geographical setting [46–48]. In South Africa, the monthly governmental cash transfers from the Child Support Grant (CSG) given to primary caregivers of children (≤18 years) have improved household access to food [49]. Moreover, about 80% of all the AGYW in HPTN-068 trial were CSG beneficiaries at baseline while about half of them received both CSG and HPTN-068 cash [34]. The HPTN-068 money given to the households was also primarily used to buy food [34]. It is plausible that being able to access multiple sources of supplemental income alleviated some stresses related to food acquisition in the study households and helped buffer adverse consequences on AGYW’s mental health symptoms [50].

Other plausible explanations for the null association identified between hunger trajectories and mental health symptoms in the AGYW include parenting practices, which can mediate this relationship [32]. Notably, most of the participants reported a supportive family environment, which may have buffered the negative impact of hunger on their mental health symptoms. Peer support is another strong protective factor for mental health [7]. We found that AGYW from households with no hunger and marginal hunger reported similar number of friends at baseline (Table 1). Other HPTN-068 studies have reported that AGYW had strong peer support systems and those from the cash transfer arm often used the money to buy gifts and food for their friends in the control arm [51]. The AGYW also had high school attendance rates [34], and reported favorable school environments. It is possible that these experiences and the presence of school feeding programs had a strong protective effect on their mental health which buffered against any harmful impacts of marginal hunger.

Hunger trajectories were not associated with hope in this study. Our study participants reported high levels of hopefulness (median score was 45, scale range 12-48), similar to findings from other studies in South Africa [7, 52]. Hope is a construct that reflects psychological strength [41]. The high levels of hopefulness reported by AGYW across all waves may have buffered against the potential psychological distress arising from any marginal hunger experience. However, it is also plausible that the Hope scale used in this study did not adequately measure the construct as intended. There was very little variation in hope scores with the same median score reported in both waves 3 and 4, suggesting a possible ceiling effect. Minimal variation in hope scores was also reported at baseline in a prior HPTN-068 study [41]. Thus, this scale may not be able to distinguish variation in hope at the high end of the spectrum over time. Another potential explanation may be that in this particular environmental and cultural setting, adolescents may perceive the concept of hope differently, which needs to be further investigated [52, 53].

The relatively short follow-up time and self-reported nature of anxiety and depressive symptoms and hopefulness outcomes prompts a cautious interpretation of our findings. Mental health symptoms may take time to manifest, and it is possible that the short follow-up period in this study did not allow for the adequate capture of incident symptomatology. Anxiety and depressive symptomology were not confirmed with clinical diagnosis. Considerable stigma exists around openly discussing mental health conditions in sub-Saharan Africa [54], so the findings could have been subject to social desirability bias [3]. We explored the sensitivity of our results to our definition of self-reported outcomes, using different cut-off scores in the RCMAS and CDI scales (Supplementary Materials Table S9-S12). These sensitivity analyses produced similar null findings, alleviating some concern around misclassification of these mental health symptoms.

A similar concern exists around the hunger measure, which was assessed at the household level and self-reported by caregivers. Household hunger may not accurately reflect the individual-level hunger of the AGYW. However, in additional analyses, we found that AGYW also self-reported increasingly lower levels of “worry” regarding their household food situation over time during the study period. Also, the HHS assesses a severe indicator of HFI, i.e., skipped meals and hunger in the last 30 days. To complement this, we included a measure of dietary diversity to gain an insight into their diet quality and found overall, households had high dietary diversity.

Finally, this study used data from school going AGYW participating in a cash transfer trial. The study participants were school-motivated, and their school attendance was much higher than generally reported in South Africa [34]. It is possible that AGYW who were not attending school would have different exposures such as no access to school feeding programs, and thus worse hunger experiences, and perceived limited opportunities for the future which can affect their mental health symptoms [41]. Therefore, these findings may have limited generalizability to AGYW who may not be attending school or those residing outside of rural Agincourt settings.

## Conclusion

This longitudinal study contributes to the scarce literature examining the association between hunger trajectories and mental health symptoms in AGYW in rural South Africa. The findings suggest a lack of association between hunger and mental health symptoms. It is possible that in this sample of AGYW, the low prevalence of moderate-severe hunger in households, access to cash transfer safety nets, high school attendance rates, and having a strong peer and family support system may have buffered against any adverse consequences of marginal hunger on their mental health. Given the recent evidence of declining prevalence of hunger in South Africa, future studies should employ other experience-based scales such as the Food Insecurity Experience Scale to monitor the full spectrum of HFI experience beyond hunger. Considering the various governmental, cultural, and social systems that may have protected these AGYW and built resilience to hunger, it is crucial to identify these actionable factors to inform the design of policies to promote adolescent and young adult health.

## Data Availability

The data archive is held at Fred Hutch Cancer Center, Seattle, WA. Requests can be sent to. The HPTN-068 data now publicly available here: https://dataverse.harvard.edu/dataverse/HPTN068

## Statements and Declarations

### Ethical Approval

This current study was deemed “Not a Human Subjects Research” by Indiana University Bloomington since we used de-identified data for all analysis. The HPTN-068 trial was approved by the Institutional Review Board of the University of North Carolina at Chapel Hill, the University of the Witwatersrand Human Research Ethics Committee, and the Mpumalanga Province Health Research and Ethics Committee. The trial is registered at clinicaltrial.gov (NCT01233531). Each young women (> 18 years) and her parents or caregivers provided written informed consent prior to HPTN-068 trial start. For AGYW under 18 years, written assent was obtained.

## Acknowledgements

The SAMRC/Wits Rural Public Health and Health Transitions Research Unit and Agincourt Health and socio-Demographic Surveillance System, a node of the South African Population Research Infrastructure Network (SAPRIN), is supported by the Department of Science and Innovation, the University of the Witwatersrand, and the Medical Research Council, South Africa, and previously the Wellcome Trust, UK (grants 058893/Z/99/A; 069683/Z/02/Z; 085477/Z/08/Z; 085477/B/08/Z).

## Author Contributions

Rishika Chakraborty: Conceptualization, Data Curation, Methodology, Formal Analysis, Writing-original draft, Writing-review and editing

Molly Rosenberg: Conceptualization, Data Curation, Methodology, Supervision, Writing-review and editing

M. Margaret Weigel: Conceptualization, Supervision, Writing-review and editing

Audrey Pettifor: Writing-review and editing, Funding Acquisition

Kathleen Kahn: Writing-review and editing

F. Xavier Gómez-Olivé: Writing-review and editing

## References

1. WHO. Mental health of adolescents 2021 [cited 2023 09/24/23]. Available from: https://www.who.int/news-room/fact-sheets/detail/adolescent-mental-health.

2. Solmi M, Radua J, Olivola M, Croce E, Soardo L, Salazar de Pablo G, et al. Age at onset of mental disorders worldwide: large-scale meta-analysis of 192 epidemiological studies. Molecular psychiatry. 2022;27(1):281–95. doi: 10.1038/s41380-021-01161-7.

3. Saade S, Lamarche AP, Khalaf T, Makke S, Legg A. What barriers could impede access to mental health services for children and adolescents in Africa? A scoping review. BMC Health Services Research. 2023;23(1):348. doi: 10.1186/s12913-023-09294-x.

4. Bundy DAP, Silva Nd, Horton S, Patton GC, Schultz L, Jamison DT. Child and Adolescent Health and Development: Realizing Neglected Potential. Disease Control Priorities, Third Edition (Volume 8): Child and Adolescent Health and Development. p. 1–24.

5. Jörns-Presentati A, Napp A-K, Dessauvagie AS, Stein DJ, Jonker D, Breet E, et al. The prevalence of mental health problems in sub-Saharan adolescents: A systematic review. Plos one. 2021;16(5):e0251689.

6. Das-Munshi J, Lund C, Mathews C, Clark C, Rothon C, Stansfeld S. Mental health inequalities in adolescents growing up in post-apartheid South Africa: cross-sectional survey, SHaW study. PloS one. 2016;11(5):e0154478.

7. Cheng Y, Li X, Lou C, Sonenstein FL, Kalamar A, Jejeebhoy S, et al. The Association Between Social Support and Mental Health Among Vulnerable Adolescents in Five Cities: Findings From the Study of the Well-Being of Adolescents in Vulnerable Environments. Journal of Adolescent Health. 2014;55(6, Supplement):S31–S8. doi: 10.1016/j.jadohealth.2014.08.020.

8. Marlow M, Skeen S, Grieve CM, Carvajal-Velez L, Åhs JW, Kohrt BA, et al. Detecting depression and anxiety among adolescents in South Africa: validity of the isiXhosa patient health Questionnaire-9 and generalized anxiety Disorder-7. Journal of Adolescent Health. 2023;72(1):S52–S60.

9. Burke MP, Martini LH, Çayır E, Hartline-Grafton HL, Meade RL. Severity of household food insecurity is positively associated with mental disorders among children and adolescents in the United States. The Journal of nutrition. 2016;146(10):2019–26.

10. Cain KS, Meyer SC, Cummer E, Patel KK, Casacchia NJ, Montez K, et al. Association of Food Insecurity with Mental Health Outcomes in Parents and Children. Academic Pediatrics. 2022;22(7):1105–14. doi: 10.1016/j.acap.2022.04.010.

11. Romo M, Abril-Ulloa V, Kelvin E, Romo ML, Kelvin EA. The relationship between hunger and mental health outcomes among school-going Ecuadorian adolescents. Social Psychiatry & Psychiatric Epidemiology. 2016;51(6):827–37. doi: 10.1007/s00127-016-1204-9.

12. Bickel G, Mark N, Cristofer P, William H, John C. Guide to Measuring Household Food Security, Revised March 2000. 2000.

13. van den Berg L, Walsh CM. Household food insecurity in South Africa from 1999 to 2021: a metrics perspective. Public Health Nutr. 2023:1–17. Epub 20230929. doi: 10.1017/s1368980023001878. PubMed PMID: 37771235.

14. May J, Witten C, Lake L. South African Child Gauge 2020. Cape Town: Children’s Institute, University of Cape Town. 2020.

15. Hayes J, Carvajal-Velez L, Hijazi Z, Ahs JW, Doraiswamy PM, El Azzouzi FA, et al. You can’t manage what you do not measure-Why adolescent mental health monitoring matters. Journal of Adolescent Health. 2023;72(1):S7–S8.

16. Weaver LJ, Hadley C. Moving beyond hunger and nutrition: a systematic review of the evidence linking food insecurity and mental health in developing countries. Ecology of Food and Nutrition. 2009;48(4):263–84.

17. Trudell JP, Burnet ML, Ziegler BR, Luginaah I. The impact of food insecurity on mental health in Africa: A systematic review. Social Science & Medicine. 2021;278:113953.

18. Jebena MG, Lindstrom D, Belachew T, Hadley C, Lachat C, Verstraeten R, et al. Food Insecurity and Common Mental Disorders among Ethiopian Youth: Structural Equation Modeling. PLoS One. 2016;11(11):e0165931. doi: 10.1371/journal.pone.0165931.

19. Nyundo A, Manu A, Regan M, Ismail A, Chukwu A, Dessie Y, et al. Factors associated with depressive symptoms and suicidal ideation and behaviours amongst sub-Saharan African adolescents aged 10-19 years: cross-sectional study. Trop Med Int Health. 2020;25(1):54–69. doi: 10.1111/tmi.13336.

20. McRell AS, Fram MS, Frongillo EA. Adolescent-Reported Household Food Insecurity and Adolescents’ Poor Mental and Physical Health and Food Insufficiency in Kenya. Curr Dev Nutr. 2022;6(8):nzac117. doi: 10.1093/cdn/nzac117.

21. Seidu A-A, Ahinkorah BO, Dadzie LK, Ameyaw EK, Budu E. Analysis of risk and protective factors for psychosocial distress among in-school adolescents in Tanzania. Journal of Public Health. 2021;29(4):765–73.

22. Shayo FK, Lawala PS. Does food insecurity link to suicidal behaviors among in-school adolescents? Findings from the low-income country of sub-Saharan Africa. BMC psychiatry. 2019;19(1):1–8.

23. Tomita A, Cuadros DF, Mabhaudhi T, Sartorius B, Ncama BP, Dangour AD, et al. Spatial clustering of food insecurity and its association with depression: a geospatial analysis of nationally representative South African data, 2008–2015. Scientific Reports. 2020;10(1):13771. doi: 10.1038/s41598-020-70647-1.

24. Pappin M, Marais L, Sharp C, Lenka M, Cloete J, Skinner D, et al. Socio-economic status and socio-emotional health of orphans in South Africa. Journal of community health. 2015;40(1):92–102.

25. Cluver L, Orkin M. Cumulative risk and AIDS-orphanhood: Interactions of stigma, bullying and poverty on child mental health in South Africa. Social science & medicine. 2009;69(8):1186–93.

26. Cluver L, Gardner F, Operario D. Poverty and psychological health among AIDS-orphaned children in Cape Town, South Africa. AIDS Care. 2009;21(6):732–41.

27. Cluver L, Orkin M, Boyes ME, Sherr L. Child and adolescent suicide attempts, suicidal behavior, and adverse childhood experiences in South Africa: a prospective study. Journal of Adolescent Health. 2015;57(1):52–9.

28. Du Toit S, Haag K, Skeen S, Sherr L, Orkin M, Rudgard WE, et al. Accelerating progress towards improved mental health and healthy behaviours in adolescents living in adversity: findings from a longitudinal study in South Africa. Psychol Health Med. 2022:1–13. Epub 20220808. doi: 10.1080/13548506.2022.2108081. PubMed PMID: 35941826.

29. Fram MS, Frongillo EA, Jones SJ, Williams RC, Burke MP, DeLoach KP, et al. Children are aware of food insecurity and take responsibility for managing food resources. J Nutr. 2011;141(6):1114–9. Epub 2011/04/29. doi: 10.3945/jn.110.135988. PubMed PMID: 21525257.

30. Bernal J, Frongillo EA, Jaffe K. Food insecurity of children and shame of others knowing they are without food. Journal of Hunger & Environmental Nutrition. 2016;11(2):180–94.

31. Wu H, Gu Z, Zeng L, Guo T. Do Global Adolescents With Food Insecurity Feel Lonely? Frontiers in Public Health. 2022;10. doi: 10.3389/fpubh.2022.820444.

32. Engel K. Parenting Stress in Households Experiencing Food Insecurity: Mental Health as a Mediator? Maternal and Child Health Journal. 2025;29(9):1244–52. doi: 10.1007/s10995-025-04131-5.

33. Bergmans RS, Malecki KM. The association of dietary inflammatory potential with depression and mental well-being among U.S. adults. Preventive medicine. 2017;99:313–9. doi: 10.1016/j.ypmed.2017.03.016.

34. Pettifor A, MacPhail C, Hughes JP, Selin A, Wang J, Gómez-Olivé FX, et al. The effect of a conditional cash transfer on HIV incidence in young women in rural South Africa (HPTN 068): a phase 3, randomised controlled trial. The Lancet Global Health. 2016;4(12):e978–e88.

35. Ballard T, Coates J, Swindale A, Deitchler M. Household hunger scale: indicator definition and measurement guide. Washington, DC: Food and nutrition technical assistance II project, FHI. 2011;360:23.

36. Nagin DS. Analyzing developmental trajectories: a semiparametric, group-based approach. Psychological methods. 1999;4(2):139.

37. Reynolds CR, Richmond BO. What i think and feel: A revised measure of children’s manifest anxiety. Journal of abnormal child psychology. 1978;6(2):271–80. doi: 10.1007/BF00919131.

38. Steventon Roberts KJ, Smith C, Cluver L, Toska E, Jochim J, Wittesaele C, et al. Adolescent mothers and their children affected by HIV—An exploration of maternal mental health, and child cognitive development. PLOS ONE. 2022;17(10):e0275805. doi: 10.1371/journal.pone.0275805.

39. Kovacs M. Children’s depression inventory (CDI and CDI 2). The encyclopedia of clinical psychology. 2014:1–5.

40. Goin DE, Pearson RM, Craske MG, Stein A, Pettifor A, Lippman SA, et al. Depression and Incident HIV in Adolescent Girls and Young Women in HIV Prevention Trials Network 068: Targets for Prevention and Mediating Factors. Am J Epidemiol. 2020;189(5):422–32. doi: 10.1093/aje/kwz238. PubMed PMID: 31667490; PubMed Central PMCID: PMCPMC7306677.

41. Abler L, Hill L, Maman S, DeVellis R, Twine R, Kahn K, et al. Hope Matters: Developing and Validating a Measure of Future Expectations Among Young Women in a High HIV Prevalence Setting in Rural South Africa (HPTN 068). AIDS Behav. 2017;21(7):2156–66. doi: 10.1007/s10461-016-1523-6.

42. Zou G. A modified poisson regression approach to prospective studies with binary data. American Journal of Epidemiology. 2004;159(7):702–6.

43. Bachman DeSilva M, Skalicky A, Beard J, Cakwe M, Zhuwau T, Simon J. Longitudinal evaluation of the psychosocial well-being of recent orphans compared with non-orphans in a school-attending cohort in KwaZulu-Natal, South Africa. International Journal of Mental Health Promotion. 2012;14(3):162–82. doi: 10.1080/14623730.2012.733600.

44. Paquin V, Muckle G, Bolanis D, Courtemanche Y, Castellanos-Ryan N, Boivin M, et al. Longitudinal Trajectories of Food Insecurity in Childhood and Their Associations With Mental Health and Functioning in Adolescence. JAMA network open. 2021;4(12):e2140085–e.

45. Zilanawala A, Pilkauskas NV. Material hardship and child socioemotional behaviors: Differences by types of hardship, timing, and duration. Children and Youth Services Review. 2012;34(4):814–25. doi: 10.1016/j.childyouth.2012.01.008.

46. Rusere F, Hunter L, Collinson M, Twine W. Patterns and trends in household food security in rural Mpumalanga Province, South Africa. Development Southern Africa. 2023:1–19. doi: 10.1080/0376835X.2023.2257737.

47. Yu X, Gill A, Chakraborty R, Kabudula CW, Wagner RG, Bassil DT, et al. Mid-life household food insecurity and subsequent memory function and rate of decline in rural South Africa, 2004-2022. Neuroepidemiology. 2024. PubMed PMID: 38857577.

48. Statistics South Africa. General Household Survey. South Africa: 2021 April 11th 2022. Report No.: Contract No.: Report.

49. DSD, SASSA, UNICEF. Child Support Grant Evaluation 2010: Qualitative Research Report. Pretoria: UNICEF South Africa. 2011.

50. Bhushan NL, Madson G, Kelly NK, Kahn K, Gomez-Olive FX, Aiello AE, et al. The relationship between household economic shocks, depression, and elevated stress-responsive biomarkers among adolescent girls and young women in rural South Africa (HPTN 068). Journal of Affective Disorders. 2025;391:119924. doi: 10.1016/j.jad.2025.119924.

51. Ndimande-Khoza MN, Scorgie F, Delany-Moretlwe S, Selin A, Twine R, Kahn K, et al. The impact of conditional cash transfers for HIV prevention on peer relationships: perspectives from female recipients and non-recipients in HPTN 068. BMC Public Health. 2022;22(1):2230. doi: 10.1186/s12889-022-14529-3.

52. Guse T, Vermaak Y. Hope, Psychosocial Well-Being and Socioeconomic Status Among a Group of South African Adolescents. Journal of Psychology in Africa. 2011;21(4):527–33. doi: 10.1080/14330237.2011.10820493.

53. Allman M, Penner F, Hernandez Ortiz J, Marais L, Rani K, Lenka M, et al. Hope and mental health problems among orphans and vulnerable children in South Africa. AIDS Care. 2023;35(2):198–204. doi: 10.1080/09540121.2022.2104795.

54. Nxumalo CT, McHunu GG. Exploring the stigma related experiences of family members of persons with mental illness in a selected community in the iLembe district, KwaZulu-Natal. Health SA Gesondheid. 2017;22:202–12. doi: 10.1016/j.hsag.2017.02.002.

